# Long Term Health Consequences of COVID-19 in Hospitalized Patients from North India: A follow up study of upto 12 months

**DOI:** 10.1101/2021.06.21.21258543

**Authors:** Sandeep Budhiraja, Mona Aggarwal, Rebecca Wig, Akansha Tyagi, RS Mishra, Monica Mahajan, Jay Kirtani, Rommel Tickoo, Supriya Bali, Arun Dewan, Ritesh Aggarwal, Prashant Saxena, Namrita Singh, Ashok Kumar, I. M. Chugh, Pankaj Aneja, Sanjay Dhall, Vandana Boobna, Vineet Arora, Sujeet Jha, Abhaya Indrayan

## Abstract

**Background:** Long-COVID syndrome is now a real and pressing public health concern. We cannot reliably predict who will recover quickly or suffer with mild debilitating long COVID-19 symptoms or battle life-threatening complications. In order to address some of these questions, we studied the presence of symptoms and various correlates in COVID-19 patients who were discharged from hospital, 3 months and up to 12 months after acute COVID-19 illness.

**Methods:** This is an observational follow-up study of RT-PCR confirmed COVID-19 patients admitted at 3 hospitals in north India between April – August 2020. Patients were interviewed telephonically using a questionnaire regarding the post-COVID symptoms. The first tele-calling was done in the month of September 2020, which corresponded to 4-16 weeks after disease onset. All those who reported presence of long COVID symptoms, were followed-up with a second call, in the month of March 2021, corresponding to around 9-12 months after the onset of disease.

**Results:** Of 990 patients who responded to the first call, 615 (62.2%) had mild illness, 227 (22.9%) had moderate and 148 (15.0%) had severe COVID-19 illness at the time of admission. Nearly 40% (399) of these 990 patients reported at least one symptom at that time. Of these 399 long-COVID patients, 311 (almost 78%) responded to the second follow-up. Nearly 8% reported ongoing symptomatic COVID, lasting 1-3 months and 32% patients having post-COVID phase with symptoms lasting 3-12 months. Nearly 11% patients continued to have at least one symptom even at the time of the second interview (9-12 months after the disease onset). Overall, we observed Long-COVID in almost 40% of our study group. Incidence of the symptoms in both the follow-ups remained almost same across age-groups, gender, severity of illness at admission and presence of comorbidity, with no significant association with any of them. Most common symptoms experienced in long COVID phase in our cohort were fatigue, myalgia, neuro-psychiatric symptoms like depression, anxiety, “brain fog” and sleep disorder, and breathlessness. Fatigue was found to be significantly more often reported in the elderly population and in those patients who had a severe COVID-19 illness at the time of admission. Persistence of breathlessness was also reported significantly more often in those who had severe disease at the onset. The overall median duration of long COVID symptoms was 16.9 weeks with inter-quartile range of 12.4 to 35.6 weeks. The duration of symptom resolution was not associated with age, gender or comorbidity but was significantly associated with severity of illness at the time of admission (P=0.006).

**Conclusions:** Long-COVID is now being recognized as a new disease entity, which includes a constellation of symptoms. Long-COVID was in almost 40% of our study group with no correlation to age, gender, comorbidities or to the disease severity. The duration of symptom resolution was significantly associated with severity of illness at the time of admission (P = 0.006). In our study, all patients reported minor symptoms such as fatigue, myalgia, neuro-psychiatric symptoms like depression, anxiety, “brain fog” and sleep disorder and persistence of breathlessness. Severe organ damage was not reported by our subjects. This might be the longest post-COVID follow-up study on a sample of nearly 1000 cases from India.

## BACKGROUND

The focus in the initial stage of COVID-19 pandemic was on detecting and treating COVID-19 related symptoms and complication. Subsequent data emerging from all over the world indicates the prolonged impact of COVID-19 infection on health of cases who recovered from acute illness and discharged from hospital. These lingering symptoms are a major concern for quality of life of recovered cases of acute COVID-19 illness as return to baseline health is delayed.

According to the guidelines published by The National Institute for Health and Care Excellence (NICE) [1], the three phases following infection consistent with COVID-19 are acute phase (for up to 4 weeks following the onset of illness), ongoing symptomatic phase (from 4 to 12 weeks following the onset of illness), and post-COVID-19 phase (symptoms that develop during or after COVID-19, and continue for ≥ 12 weeks, not explained by an alternative diagnosis). The long COVID-19 phase (signs and symptoms that continue or develop after acute COVID-19) includes both ongoing symptomatic COVID-19 (from 4 to 12 weeks) and post-COVID-19 syndrome (12 weeks or more).

The long-COVID-19 is affecting a growing number of COVID-19 patients and has become a major cause of increasing burden on healthcare system. Long-term follow-up studies on the prolonged symptoms have been published from many countries with longest follow up of around 6-9 months till date [2, 3, 4, 5], but the adequate knowledge is lacking on persistence for higher duration and on specific symptoms, duration for recovery and association with any comorbidity, severity of acute phase of COVID-19 illness and age of the patient, particularly for the patients in India.

We assessed persistent symptoms up to one year in patients who were discharged from the hospital after recovery from acute phase of COVID-19 in India, with an aim to describe the consequences of long COVID and to identify potential factors associated with those symptoms. This can help in assessing the need and type of long-term follow-up and rehabilitation programs required for patients hospitalised for COVID -19.

## METHODS

This is an observational follow-up study of COVID-19 patients who were admitted at 3 hospitals in northern India between April–August 2020. All the patients were contacted twice over the telephone for follow-up. First follow-up was done in September 2020 (4 weeks to 16 weeks from the onset of disease) and those agreeing were interviewed using a questionnaire. This captured details of each patient during hospitalization and their status post-discharge. Nobody was personally examined. Figure 1 shows the CONSORT – like diagram for the present study.

**Figure 1:**
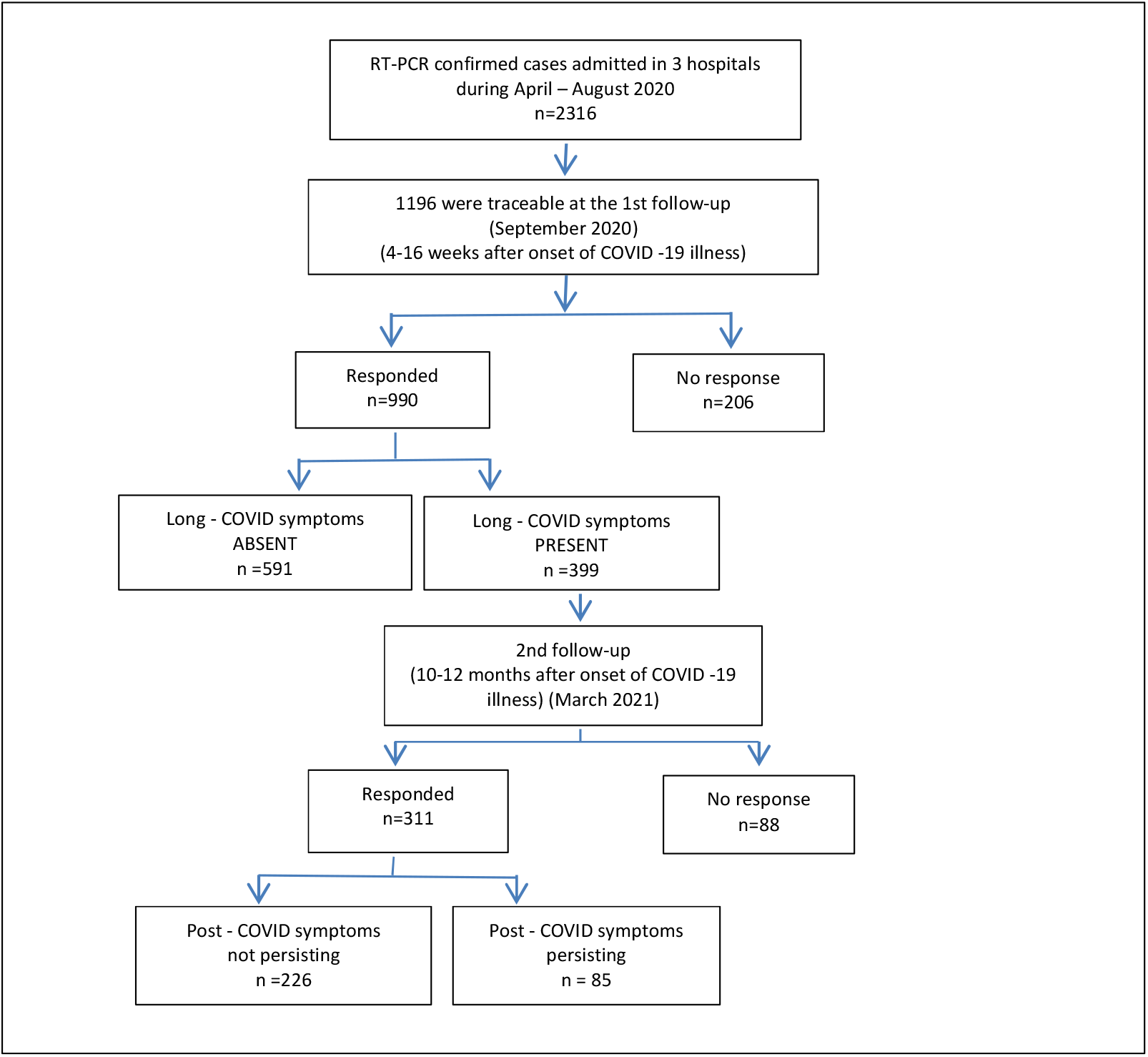
CONSORT Diagram.

In March 2021, second follow-up with those who reported symptoms during the first follow-up was done using the questionnaire to capture the status and profile of prolonged symptoms of long COVID.

The pre discharge and discharge data was extracted from the hospital’s Computerized Patient Record System (CPRS). Important data points such as those required for assessing severity were captured. The cases were divided into mild, moderate, and severe categories of COVID-19 illness, based on the Government of India protocol [6]. This included considerations such as ICU admission and the use of ventilator.

The initial analysis of the data was done by identifying the prevalence of various symptoms such as fatigue weakness, breathlessness in different age-groups, gender, and disease-severity groups. The association of presence of symptoms with these characteristics was checked for statistical significance with chi-square test or with Fisher exact test in case the number of subjects in any category was too small (<5). The significance of difference between mean durations of symptom among different age-groups, and severity groups, was checked with ANOVA F-test, and the difference between gender was checked with Student t-test. The level of significance was fixed at 5% and SPSS21 was used for calculations.

## RESULTS

A total of 2316 RT-PCR confirmed COVID-19 patients were admitted to the 3 hospitals under study during April – August 2020. Of these, 1196 (51.6%) were traceable and 990 (42.7%) responded to our first follow-up. This was done 4-16 weeks from the onset of COVID-19 illness, with an average of 9 weeks. As many as 399 (40.3%) of 990 cases reported at least one symptom in the first follow-up. In the second follow-up, 311 out of 399 (77.9%) responded, and 85 (27.3%) reported that the symptoms were still persisting at the time of interview 9-12 months from the disease onset. The remaining 226 (72.7%) reported resolution of symptoms in between the two follow-ups.

The characteristics of the initial cohort of 990, and of 311 patients at the second follow-up, respectively, are in Table 1. Age of these patient ranged from 1-91 years, with nearly 60% belonging to the age-group 30-59 years and 25 % to the age >60 years during both the follow ups.

**Table 1.** Patient characteristics –interviewed at first follow-up, and second follow-up.

| Characteristics | Cases in the 1<sup>st</sup> follow-up |  | Reporting symptoms at the 1<sup>st</sup> follow-up |  | Cases in the 2nd follow-up |  |
| --- | --- | --- | --- | --- | --- | --- |
|  | n | % | N | % | n | % |
| Total cases | 990 | 100.0% | 399 | 100.0% | 311 | 100.0% |
| Age group (years) |  |  |  |  |  |  |
| ≤29 | 145 | 14.6% | 55 | 13.8% | 44 | 14.1% |
| 30-59 | 591 | 59.7% | 242 | 60.6% | 188 | 60.5% |
| 60+ | 254 | 25.7% | 102 | 25.6% | 79 | 25.4% |
| Gender |  |  |  |  |  |  |
| Female | 320 | 32.3% | 141 | 35.3% | 112 | 36.0% |
| Male | 670 | 67.7% | 258 | 64.7% | 199 | 64.0% |
| Severity at admission |  |  |  |  |  |  |
| Mild | 615 | 62.2% | 240 | 60.2% | 188 | 60.5% |
| Moderate | 227 | 22.9% | 97 | 24.3% | 75 | 24.1% |
| Severe | 148 | 14.9% | 62 | 15.5% | 48 | 15.4% |
| Pre-existing comorbidities |  |  |  |  |  |  |
| None | 621 | 62.7% | 239 | 59.9% | 166 | 53.4% |
| At least one | 369 | 37.3% | 160 | 40.1% | 145 | 46.6% |
|  | Mean | SD | Mean | SD | Mean | SD |
| Duration of hospitalization (days) | 9.5 | 6.1 | 9.6 | 6.3 | 9.7 | 6.6 |
| Duration of symptom at admission (days) | 4.9 | 4.3 | 5.9 | 5.3 | 5.1 | 4.7 |

Among 990 respondents of the first follow-up, 320 (32.3%) were female and 670 (67.7%) were male. Covid-19 illness severity at the time of hospitalization was mild in 62.2%, moderate in 22.9%, and serious in 14.9%. More than one-third (37.3%) reported at least one co-morbidity at admission – diabetes (23.7%) and hypertension (20.4%) being the most common. The average duration of hospitalization was 9.5 days (SD = 6.1 days) and they had COVID 19 symptoms for an average of 4.9 days (SD = 4.3 days) at the time of admission. These percentages and averages were similar in those interviewed at second follow-up (Table 1) and show that the followed-up cases second time were fair representative of the cases responding to the first interview. Among 311 cases responding to the second round of interview in March 2021, 112 (36.0%) were females and 199 (64.0%) were male, and 145 (46.6%) reported to have at least one co-morbidity at the time of hospitalisation.

Severity of the disease at the time of admission was significantly associated with age (P<0.001). Mild cases were relatively more in cases of 0-29 years age and serious more in cases of age >60 years or more (Table 2). As expected, pre-existing comorbidity was present in only 8.3% patients of age less than 30 years, 34.2% in patients of age 30-59 years and 61.0% in patients of age 60 years and above. The association of age with the presence of comorbidity was highly significant (P<0.001). Moderate and serious cases reported at least one comorbidity (48.9% and 45.3% respectively), significantly more often (P<0.001) than in mild cases (31.1%). Diabetes was reported more often (43.3%) in cases with moderate severity compared to mild (19.3%) and severe (25.7%) cases (P<0.001) (Figure 2). Hypertension was reported more often in cases with severe (26.4%) and moderate (24.7%) disease as compared to mild cases (17.4%) (P= 0.010). Hypothyroidism was twice more common in cases with severe illness (6.8%) compared to the cases with mild illness (3.3%).

**Table 2.**
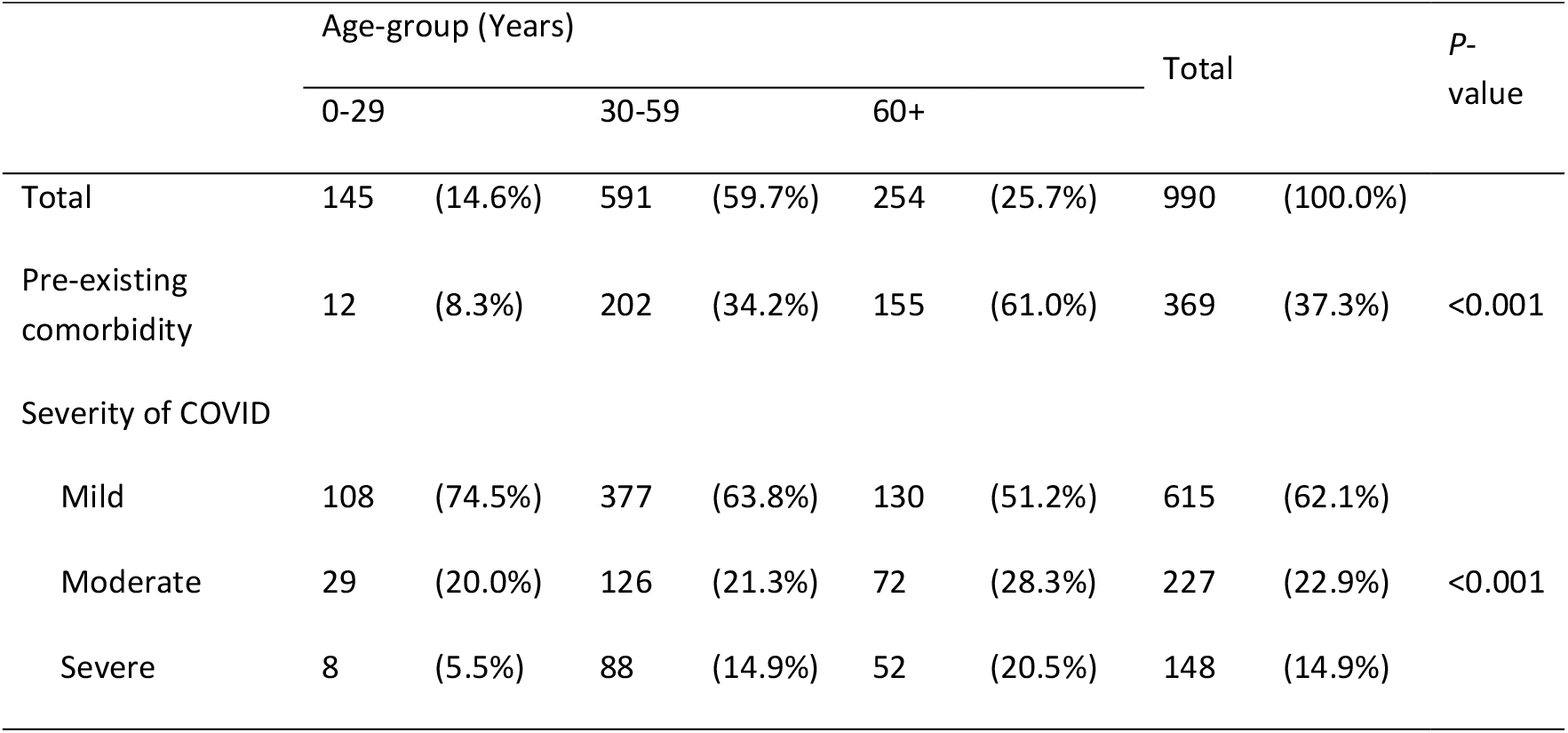
Comorbidities and severity in different age-groups.

**Figure 2.**
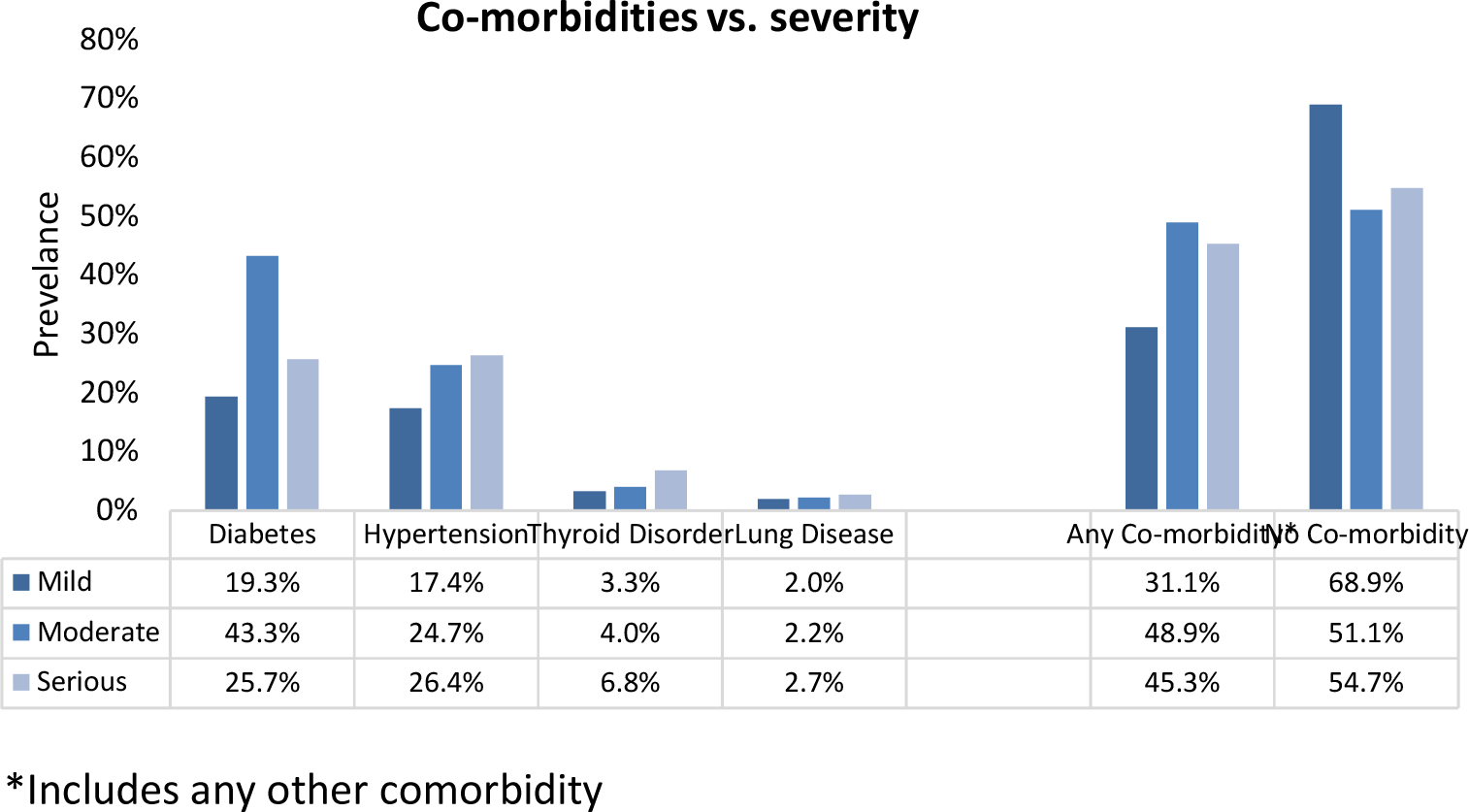
Prevalence of different co-morbidities in mild, moderate, and severe cases.

As mentioned earlier, 399 (40.3%) cases reported at least one symptom at the time of the first interview and this percentage remained nearly the same across different age-groups, gender, severity at admission, and the presence of comorbidity (Table 3). Thus the presence of symptoms at the first interview were not significantly associated with any of these characteristics (minimum P = 0.096 for gender). More than one-fourth (27.3%) cases responding to the second interview reported at least one persisting symptom and this percentage too remained fairly stable across age-groups, gender, severity at admission, and the presence of comorbidity with no significant association with any of them (minimum P = 0.289 for gender). Interestingly, we observed that average duration of symptoms prior to hospitalization was significantly more in those patients who ultimately reported long-COVID symptoms, during both interviews, with P-values being 0.001 and 0.026 for the first and second tele-interviews, respectively.

**Table 3.** Patients reporting symptoms at the first follow-up and continuing at the time of the second follow-up.

| Characteristic | Symptoms- 1<sup>st</sup> follow-up |  |  |  |  | Persisting Symptoms -2<sup>nd</sup> follow-up |  |  |  |  |
| --- | --- | --- | --- | --- | --- | --- | --- | --- | --- | --- |
|  | At least One |  | None |  | P-value* | At least One |  | None |  | P-value* |
|  | N | % | n | % |  | n | % | n | % |  |
| Total cases | 399 | 40.3% | 591 | 59.7% |  | 85 | 27.3% | 226 | 72.7% |  |
| Age group (years) |  |  |  |  |  |  |  |  |  |  |
| ≤29 | 55 | 37.9% | 90 | 62.1% | 0.801 | 12 | 27.3% | 32 | 72.7% | 0.993 |
| 30-59 | 242 | 40.9% | 349 | 59.1% |  | 51 | 27.1% | 137 | 72.9% |  |
| 60+ | 102 | 40.2% | 152 | 59.8% |  | 22 | 27.8% | 57 | 72.2% |  |
| Gender |  |  |  |  |  |  |  |  |  |  |
| Female | 141 | 44.1% | 179 | 55.9% | 0.096 | 35 | 31.3% | 77 | 68.8% | 0.289 |
| Male | 258 | 38.5% | 412 | 61.5% |  | 50 | 25.1% | 149 | 74.9% |  |
| Severity at admission |  |  |  |  |  |  |  |  |  |  |
| Mild | 240 | 39.0% | 375 | 61.0% | 0.568 | 47 | 25.0% | 141 | 75.0% | 0.464 |
| Moderate | 97 | 42.7% | 130 | 57.3% |  | 22 | 29.3% | 53 | 70.7% |  |
| Severe | 62 | 41.9% | 86 | 58.1% |  | 16 | 33.3% | 32 | 66.7% |  |
| Pre-existing comorbidities |  |  |  |  |  |  |  |  |  |  |
| None | 239 | 38.5% | 382 | 61.5% | 0.131 | 47 | 28.3% | 119 | 71.7% | 0.678 |
| At least one | 160 | 43.4% | 209 | 56.6% |  | 38 | 26.2% | 107 | 73.8% |  |
|  | Mean | SD | Mea<br>n | SD | P-value | Mean | SD | Mean | SD | P-value |
| Duration of hospitalization (days) | 9.6 | 6.3 | 9.5 | 6.1 | 0.803 | 10.7 | 9.0 | 9.3 | 5.3 | 0.092 |
| Duration of symptoms at admission (days) | 5.9 | 5.3 | 4.9 | 4.3 | 0.001 | 6.1 | 5.8 | 4.8 | 4.0 | 0.026 |

\* P-value for association between the characteristic and the presence of at least one symptom

Many patients reported multiple symptoms. Overall occurrence of long COVID-19 symptoms reported at the second follow-up also showed no significant association with age, gender, COVID-19 severity at the time of admission, and with presence of any comorbidity (minimum P = 0.245 for gender) (Table 4). Among specific symptoms, fatigue was the most common, reported by 12.5% cases followed by myalgia (9.3%). Fatigue showed significant association with age (P = 0.017), with only 1 of 44 (2.3%) in age group of less than 30 years had fatigue, which increased to 21.5% in the age-group of 60 years or more. Fatigue was also significantly associated with severity of COVID-19 illness at admission (P = 0.016). Fatigue was reported by 22.9% among cases with severe illness at admission but only by 8.5% by the cases with mild illness (Table 4). Persistence of breathlessness was also significantly reported more in patients with severe COVID (14.6%) as compared to those with mild (4.3%) and moderate (9.3%) disease. Neuro-psychiatric symptoms, such as anxiety, depression, “brain-fog”, and sleep disorder, were reported by 9.0% cases. It was significantly associated with pre-existing comorbidity (P=0.018) and not with age, gender, or severity of illness.

**Table 4.**
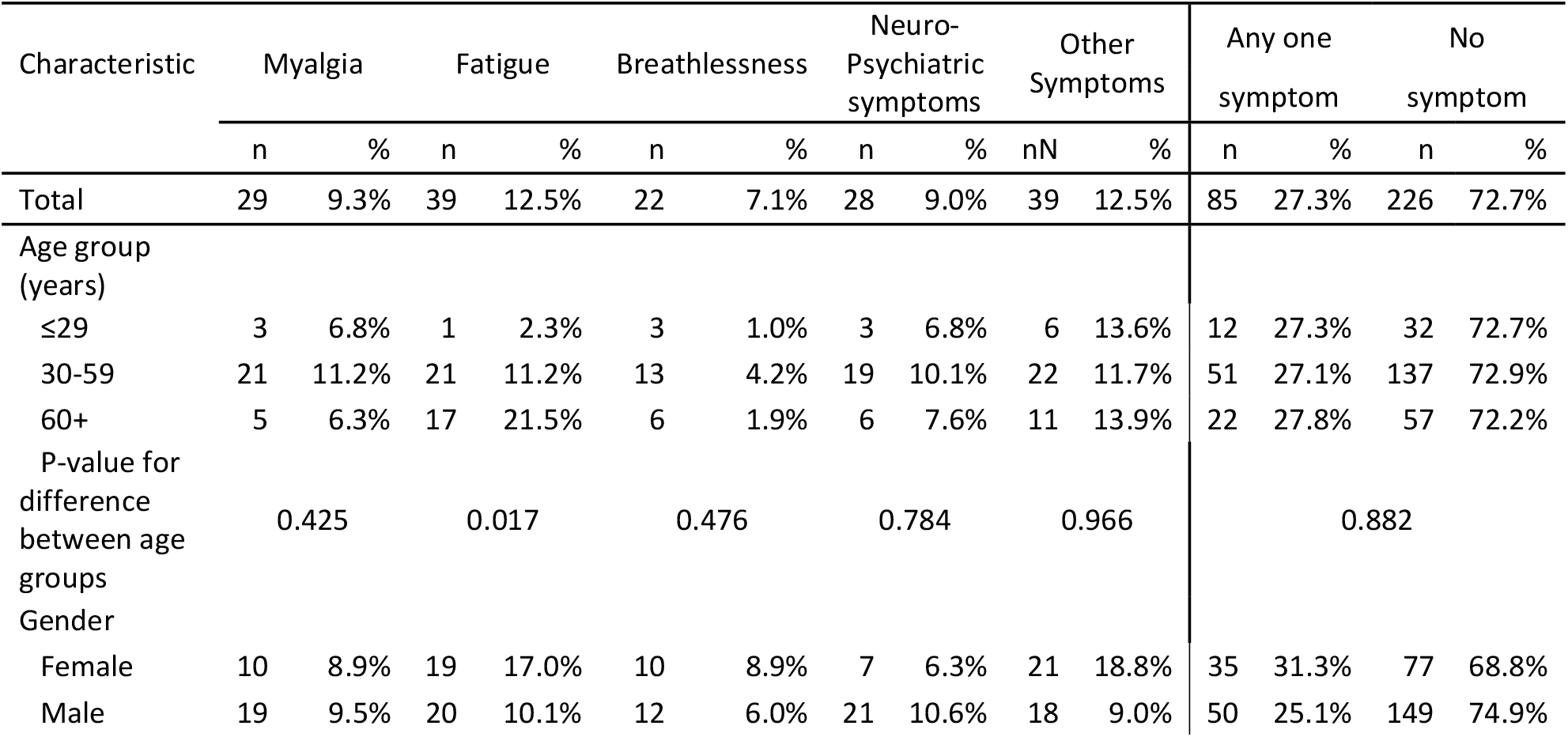
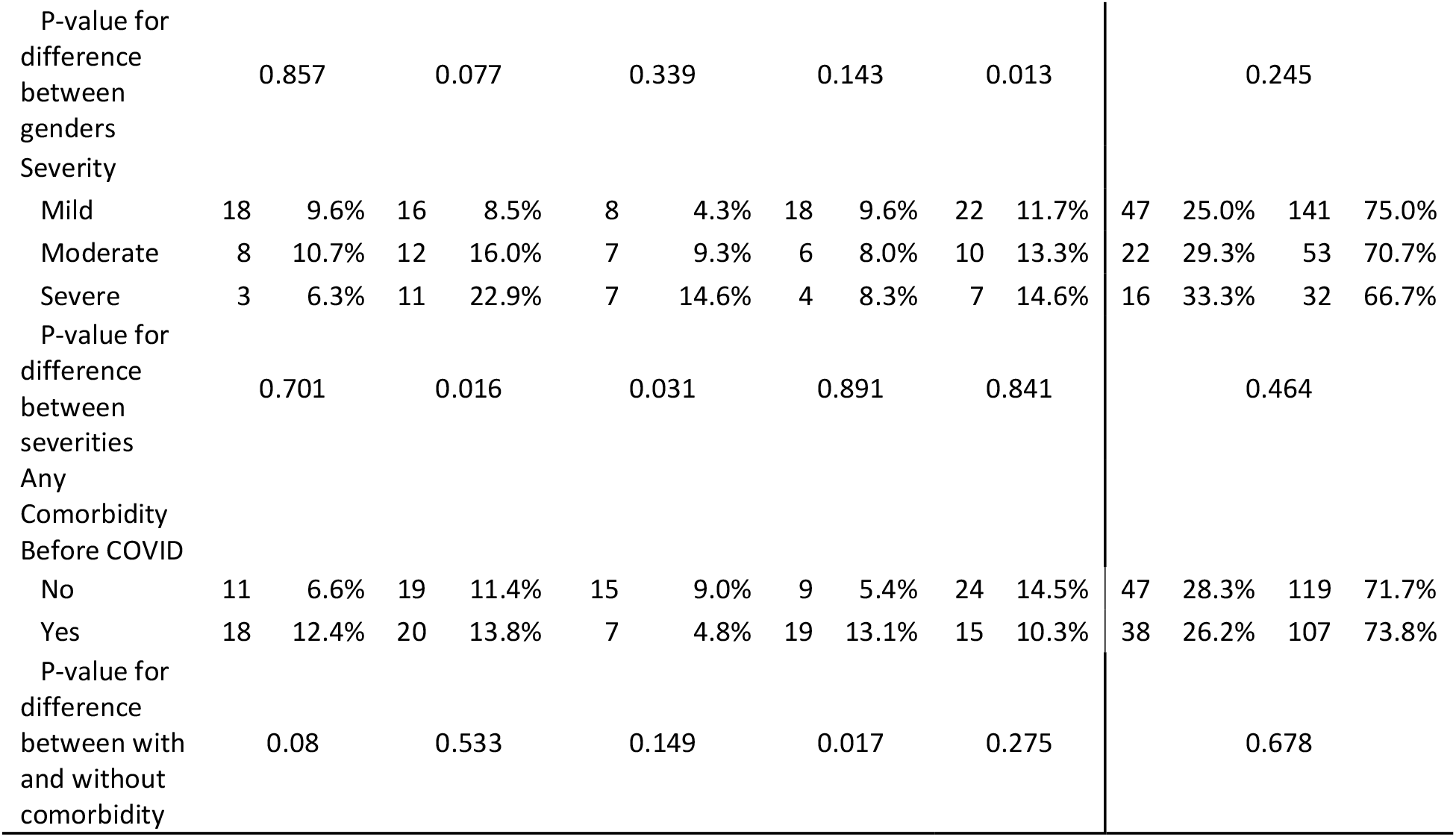
Characteristics of the patients reporting common symptoms at the time of 2^nd^ follow-up. (Percentage out of cases of that category)

From our original cohort of 990 patients, 399 reported presence of symptoms. From this group, there was eventual dropout of 88 patients during the second follow-up and 311 responded. To account for this attrition proportionate, a correction factor (1.283) was applied to reach at the percentages from the original cohort of 990 (Table 5). 62 patients (8%) reported symptoms categorized as “ongoing symptomatic phase” as per the NICE guidelines since their symptoms lasted for between 1-3 months. The number of patients reporting post-COVID symptoms were: 118 (15.3%) with symptoms lasting 3-6 months, 43 (5.5%) with symptoms lasting 6-9 months and 85 (11%) patients continued to have symptoms even at the time of interview (9-12 months from the symptom onset). So, in all, 246 (31.6%) patients reported post-COVID symptoms in our study. Long-COVID phase (that is, both ongoing symptomatic phase and the post-COVID phase) was seen in a total of almost 40% cases in our study.

**Table 5:**
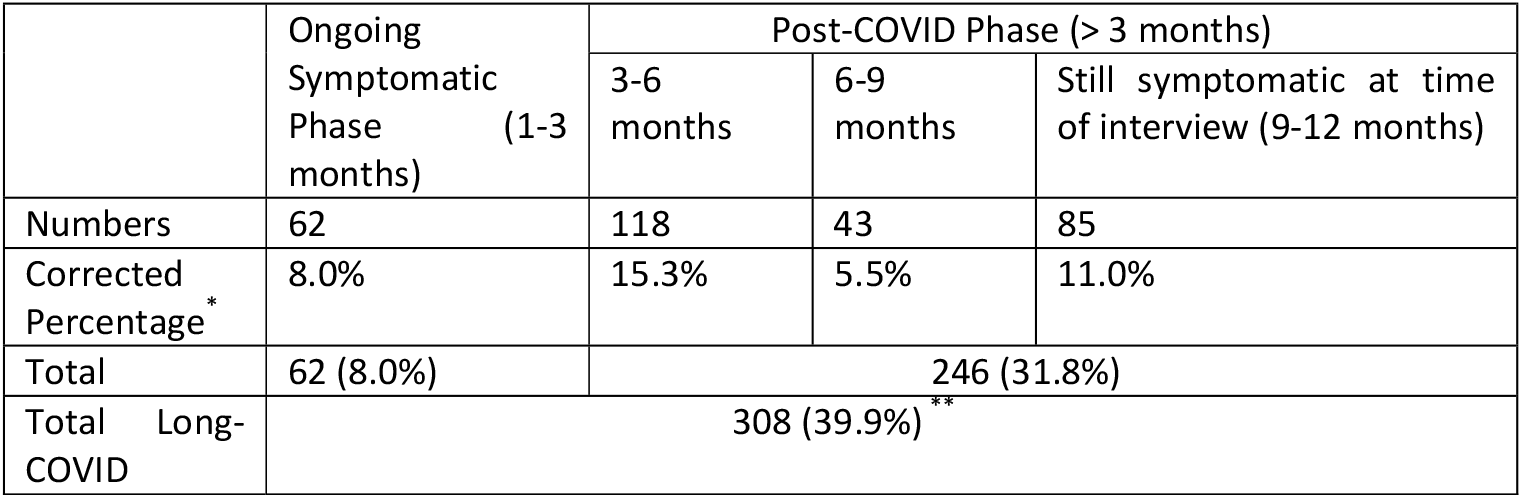
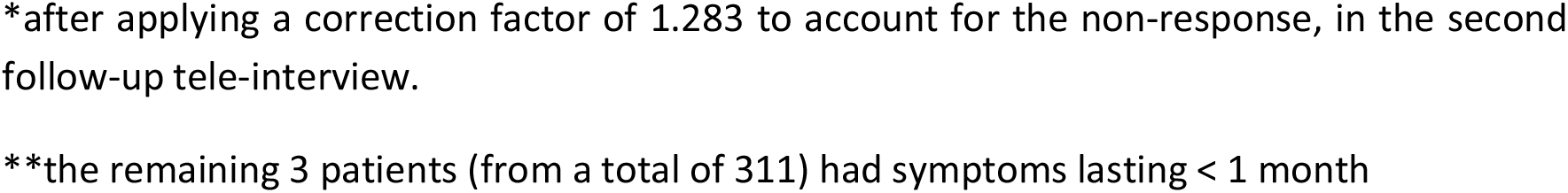
Long-COVID: Based on 2^nd^ follow-up tele-interview of 311 patients.

|  | Ongoing Symptomatic Phase (1-3 months) | Post-COVID Phase (> 3 months) |  |  |
| --- | --- | --- | --- | --- |
|  |  | 3-6 months | 6-9 months | Still symptomatic at time of interview (9-12 months) |
| Numbers | 62 | 118 | 43 | 85 |
| Corrected Percentage * | 8.0% | 15.3% | 5.5% | 11.0% |
| Total | 62 (8.0%) | 246 (31.8%) |  |  |
| Total Long-COVID | 308 (39.9%) ** |  |  |  |

\*after applying a correction factor of 1.283 to account for the non-response, in the second follow-up tele-interview.

Of 331 responding to the second interview, 214 (68.8%) reported that they were able to resume their daily routine within a month of discharge, 77 (24.8%) took 1 to 3 months, and 20 (6.4%) took 4 months or more. The differences across age-groups, gender, severity, and pre-existing comorbidity was not significant (minimum P = 0.307)

The duration of symptom resolution was not associated with age, gender, or comorbidity (minimum P = 0.573 for age) but was significantly associated with severity of illness at the time of admission (P = 0.006) (Table 6). Out of 311 cases who had long COVID-19, 188 (60.4%) had mild illness at the time of hospitalisation, 75 (24.1%) had moderate and 48 (15.4%) had severe illness. Among 188 cases with mild illness at the time of hospitalisation, 1 (0.5%) case took less than 4 weeks, 42 (22.3%) cases took 4-12 weeks and 145 (77.1%) took more than 12 weeks for symptoms to disappear completely. But among those with moderate and severe illness, 76.0% and 91.7%, respectively, took more than 12 weeks for the last symptom to disappear. The overall median duration of symptoms was 16.9 weeks with inter-quartile range of 12.4 to 35.6 weeks. This was nearly the same for each subgroup except relatively higher (20.5 weeks) for those who had serious condition at the time of admission.

**Table 6.**
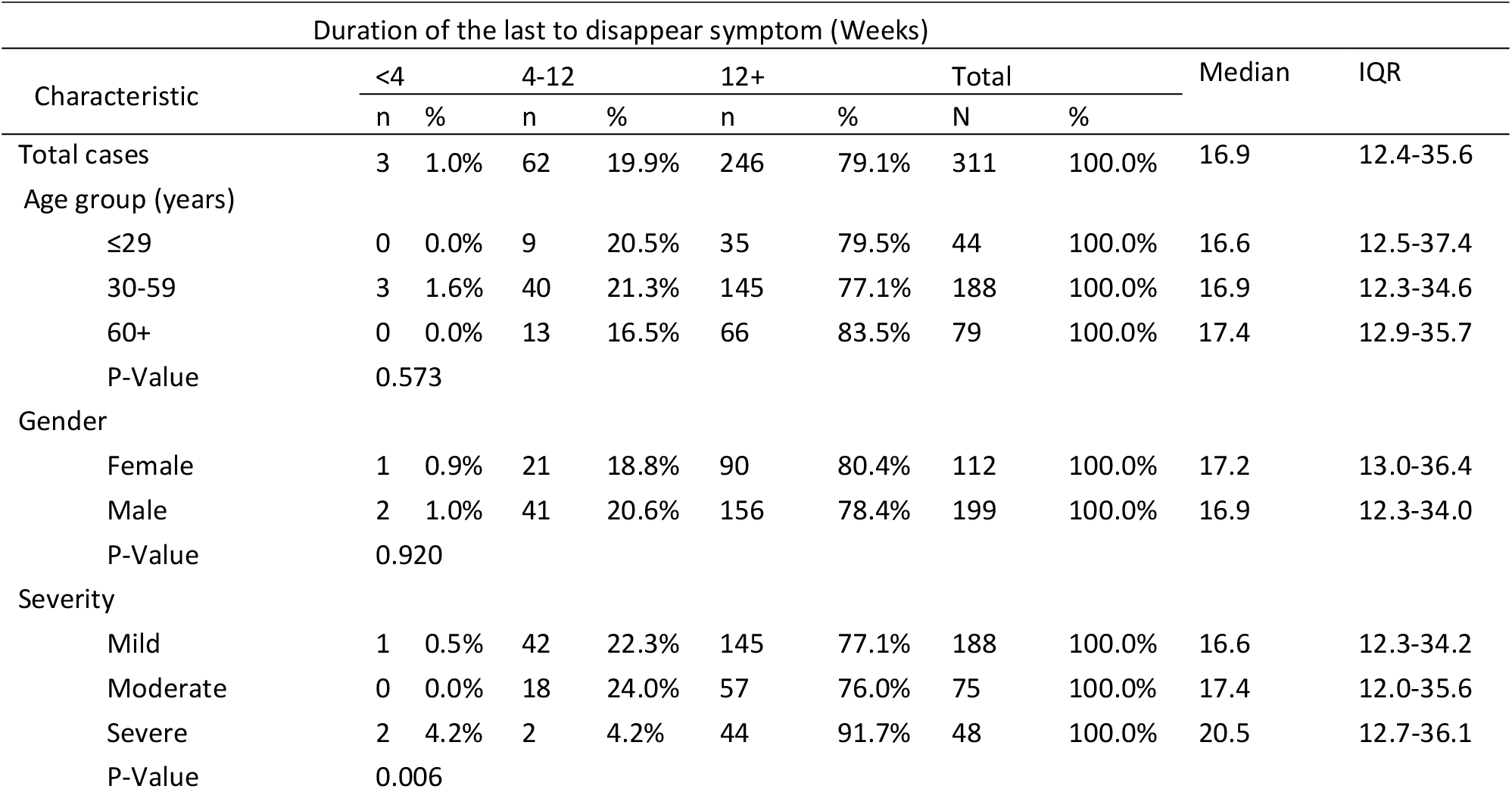
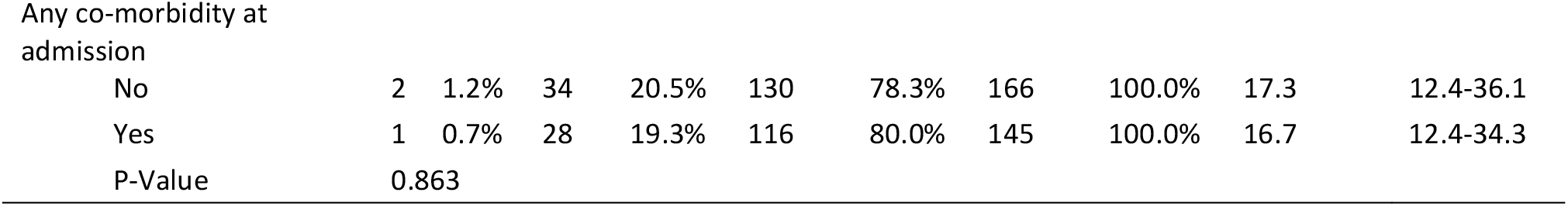
Duration of “last to disappear” symptoms.

## DISCUSSION

With the pandemic relenting in most parts of the world, the focus is shifting from the treatment modalities to long COVID-19 syndrome. This is assuming importance because a surge of discharged patients can place an additional burden on the healthcare system, patients’ families, and the society [7]. Oronsky et al. [7] from USA and Fernandes et al. [8] (from Brazil) provided a review of such symptoms. Walitt and Bartrum [9] provided a clinical primer for the expected and potential post-COVID syndrome. The frequency with which these symptoms occur is not yet well-established and varied from area to area.

This study was undertaken to shed light on the burden of prolonged symptoms faced by the hospitalized COVID-19 patients in India after discharge and their duration in a follow-up of up to one year. The present study was designed as a 2-step tele-interview, based on symptoms recall to a set of questions.

As per the prevailing Government of India guidelines during the early phase of pandemic in India (April-May 2020), all patients of RT-PCR confirmed COVID-19, needed to be admitted. Hence, hospitalized patient during our study period (April-August 2020) represented patient profile across all severity levels of COVID-19 (mild, moderate and severe). The results of this study, can therefore, be extrapolated for occurrence of Post-COVID syndrome, in mild COVID-19 patients, who were treated outside of the hospital.

In our patients, the severity of disease at admission, as expected, was found to be significantly associated with age (P < 0.001) with more severe cases in age 60 years or more. Similarly, presence of comorbidity was significantly associated with age (P < 0.001) with almost 61% patients of age 60 years or more reported at least one comorbidity. Diabetes (P < 0.001) and hypertension (P = 0.010) were significantly more prevalent in patients presenting with moderate and serious disease on admission. Interestingly, hypothyroidism was twice more common in cases with severe illness (6.8%) as compared to cases with mild illness (3.3%).

We observed, 8% patients had ongoing symptomatic phase of COVID, lasting 1-3 months, and 31.8% patients had Post-COVID phase, with symptoms lasting 3-12 months. 11% patients continued to have at least one symptom even at the time of the second interview (9-12 months from the disease onset). Overall, Long-COVID was reported in almost 40% of our study group. Occurrence of Post-COVID symptoms had no correlation to age, gender, comorbidities or to the disease severity. The duration of symptom resolution was also not associated with age, gender or comorbidity but was found to be significantly associated with severity of illness at the time of admission (P = 0.006).

Fatigue was significantly more prevalent amongst the elderly patients and in those who had severe COVID-19. Similarly, persistence of breathlessness was significantly more often reported by those patients, who had severe illness at presentation. Presence of neuro-psychiatric symptoms like depression, anxiety, “brain fog” and sleep disorders were reported in nearly 9% of cases. No patient reported any significant organ damage.

Although a significant number of patients (16.5%) continued to have some form of post COVID symptoms for as long as 6-12 months, a majority of them, had resumed their routine work much earlier. 31.2% patients took 1 month to resume daily routine while 6.4% took almost 4 months to resume their daily routine.

In a similar phone-call study in Spain [10] on 134 hospitalized COVID patients 90 days after discharge, the most frequently reported symptoms were asthenia, dyspnoea, and weight loss. Garrigues et al. [11] also conducted a telephonic study of 120 hospitalized patients in France and reported that a high proportion of individuals reported fatigue (55.0%), dyspnoea (41.7%), cough (16.7%), and chest pain (10.8%) with many reporting a worsened quality of life. In a multistate telephonic survey in the U.S., Tenforde et al. [12] reported after a telephonic interview of 294 patients that 35% had not returned to their usual state of health when interviewed 2-3 weeks after testing. They reported for patients aged 18-34 years with no chronic medical condition that almost one in four had not returned to their usual state of health by the time the interview was conducted.

The study by Townsend et al. [13] on 233 patients in Ireland, where they follow-up their patients for 6 weeks post-discharge, it was observed that a significant burden of post-viral fatigue existed in individuals with previous SARS-CoV-2 infection after the acute phase of COVID-19 illness. In a study on 89 patients in the early phase of the pandemic in China [14], the most frequently reported symptoms were cough (46.1%), fatigue (36.0%), and expectoration (24.7%), 3 weeks after discharge.

The study conducted in the Netherlands on persistent symptoms 3 months after a SARS-CoV-2 infection by Goertz et al. [15] found fatigue (92.9%), dyspnoea (89.3%), and cough (79.5%) were the most prevalent symptoms in hospitalized patients.

The long term health consequences in 1733 COVID-19 patients at an average of 6 months following discharge from hospital were evaluated by Huang et al. [3] They observed persistence of at least one symptoms in 76% cases in their cohort at the end of 6 months after disease onset. They reported fatigue (63%), sleep difficulties (26%) and anxiety or depression (23%) as the most commonly reported post COVID symptoms. They also conducted the pulmonary function test and HRCT chest in their follow up for selected patients and found that pulmonary diffusion abnormality was higher in patients with severe COVID-19 disease at acute phase.

The study by Logue et al. [4] on 177 patients with nearly 85% outpatients with mild disease, observed the persistence of long COVID-19 symptoms in 30 % cases at the single follow up done after 3-9 months (average 6 months) of COVID 19 illness. Similar to our observation, fatigue was reported the most common symptoms in this study.

Davis et al [5] conducted an international web-based survey of suspected and confirmed COVID-19 cases to characterize the symptom profile and time course in patients with long COVID, along with the impact on daily life, work and return to baseline health. Of 3762 respondents from 56 countries who completed the survey, only 8.4% were hospitalized and only 27% had a laboratory confirmed diagnosis of COVID-19. The most frequent symptoms reported after 6 months were fatigue (77.7%), post-exertional malaise (72.2%) and cognitive dysfunction (55.4%). Also, 44.7% respondents required a reduced work schedule compared to pre-illness and 22.3% were not working at the time of the survey due to their health conditions.

Daugherty et al [16] conducted a retrospective study in the United States to evaluate the excess and relative hazards for developing incident clinical sequelae after the acute phase of SARS-CoV-2 infection in adults aged 18-65 years. They had three comparator groups, matched by propensity score: a 2020 group, a historical 2019 group, and a historical group with viral lower respiratory tract illness. They observed 14% of adults less than 65 years who were infected with SARS-CoV-2 had at least one new type of clinical sequelae that required medical care after the acute phase of the illness, and this was 4.5% higher than in in the 2020 comparator group. Although older individuals with pre-existing conditions, admitted to the hospital because of COVID-19 were at highest risk, younger adults (aged < 50 years), and those with no pre-existing conditions, or those not admitted for COID-19 also had an increased risk of developing new clinical sequelae.

The pathogenesis of post-COVID syndrome is multifactorial. Prolonged inflammation has a key role in its pathogenesis and may account for some neurological complications, cognitive dysfunction and several other symptoms. Other pathogenetic mechanisms that are implicated in post-COVID syndrome include immune-mediated vascular dysfunction, thromboembolism and nervous system dysfunction [17]. The current data indicated that the overwhelming majority of patients with post-COVID syndrome have a good prognosis.

This entity of “Post-COVID Syndrome’’ needs to be explored further. It is not fully known yet what kind of patients gets this affliction, how many days such symptoms persist and with what outcome. This may vary from area to area but highlights the need for long term follow-up by a multidisciplinary team including general practitioners, physiotherapists, occupational therapists, and rehabilitation programs to improve their quality of life of these patients [18].

To just make projections on the potential burden on the health systems that might be brought about by long-COVID in India, we extrapolated our findings to the presently reported COVID-19 numbers. As on 4^th^ June 2021, India has had 2.87 crore reported cases of COVID-19. If we consider the hospitalization rate at 6% [19], then almost 17 lakhs would have required admission. If 40% of these develop long-COVID syndrome, this makes it about 7 lakh cases requiring help after discharge. Actual number of long-COVID can be much more as this condition can occur in the mild-COVID-19 patients, who were never hospitalized. This will be a very important consideration when planning for future resources and manpower, not only for India but also for the rest of the world.

### LIMITATIONS OF PRESENT STUDY

All three hospitals under study received patients from distant areas that made it difficult for the patients to visit the clinic for follow up. This was the main reason for choosing telephonic survey to conduct this study. Since based on telephonic conversations, the findings might be subject to recall bias and a full syndrome could not be studied. The interview reveals the perception and that is what matters for seeking medical help. Nonresponse at the second follow-up required that the results are adjusted.

## CONCLUSIONS

Long-COVID is now being recognized as a new disease entity, which includes a constellation of symptoms. We found that 31.8% patients had post COVID symptoms beyond 3 months, and 11% of the patients continued to have some form of symptoms for as long as 9-12 months from the onset of disease. Overall, long-COVID occurred in almost 40% of the cases. All patients with post-COVID reported only minor symptoms like fatigue, myalgia, breathlessness, depression and anxiety. Severe organ damage was not seen in our subjects. Occurrence of these symptoms had no correlation to age, gender, comorbidities or to the disease severity. However, severity of disease at the onset, was the only significant determinant of the duration of post-COVID symptoms. To the best of our knowledge, this is the longest post-COVID follow-up study from India.

## Data Availability

all data available in manuscript

## ABBREVIATIONS

Not applicable

## DECLARATIONS

### Ethics approval and consent to participate

The study titled “Long term health consequences of COVID-19 in hospitalized patients from North India: A follow up study of upto 12 months” was approved by the Institutional Ethics Committee, Devki Devi Foundation; address: service floor, office of Ethics Committee, East Block, Next to Conference Room, Max Super Speciality Hospital, Saket (A unit of Devki Devi Foundation), 2, Press Enclave Road, Saket, New Delhi – 110017 vide ref. no. BHR/RS/MSSH/DDF/SKT-2/IEC/IM/21-14, dated 18^th^ June’2021. The IEC has observed that study has been given approval by Ethics committee in the beginning on 20^th^ April’2020 vide ref. no. RS/MSSH/DDF/SKT-2/IEC/IM/20-16.

### Consent for publication

All patients admitted in these 3 hospitals gave a generic consent at the time of admission for their data to be used anonymously for research purpose. Only those who were willing were interviewed.

### Availability of data and material

Yes

### Author’s contributions

SB designed the study concept, contributed patients for the study and wrote the manuscript; MA and RW wrote the manuscript; AT prepared and executed the patient questionnaire and data collection; AI did the statistical analysis and manuscript writing; RSM, MM, JK, RT, AD, SB, RA, PS, NS, AK, IMC, PA, SD, VB and VA contributed patients for the study; SJ designed the study concept and reviewed the manuscript.

## Acknowledgements

We wish to acknowledge and thank the contribution made by Taruna Sharma, Archa Misra, Rizwan Ahmed, Shivanku, Praveen.

